# MASHA: A Multi-Agent System for Healthcare Sentiment Analysis Using AI for Migraine Detection in Arabic Tweets

**DOI:** 10.64898/2026.05.21.26352626

**Authors:** Shadia Yahya Baroud

## Abstract

Migraine detection and sentiment analysis in healthcare have become increasingly important, particularly with the rise of social media platforms like Twitter, where users often share their personal health experiences. This study presents MASHA (Multi-Agent System for Healthcare Sentiment Analysis), an artificial intelligence (AI)-driven framework that integrates multiple machine learning (ML) models for sentiment analysis of Arabic tweets related to migraines. The system leverages a multi-agent architecture to handle tasks such as data acquisition, pre-processing, model training and real-time decision-making. Key ML models, including Support Vector Machines (SVM), Naive Bayes (NB) and Logistic Regression (LR), are integrated using ensemble techniques, leading to improved classification performance.

Experiments conducted on a dataset of Arabic tweets demonstrate that MASHA outperforms traditional methods, achieving an accuracy of 90.0% and an F1-score of 89.46%. Moreover, the system’s scalability and flexibility make it suitable for real-time public health monitoring, offering valuable insights into patient experiences and public sentiment regarding healthcare services.

MASHA’s adaptability suggests its potential application for analysing other healthcare-related conditions, reinforcing the system’s scalability and broader relevance. Future work will focus on incorporating deep learning (DL) models and expanding the dataset with content from additional social media platforms.

## 1. Introduction

The integration of machine learning (ML) models is crucial in improving the performance of complex applications, particularly in the field of Natural Language Processing (NLP) (1). NLP involves the interaction between computers and human language, enabling machines to understand, interpret and generate human language (2–4). The effective integration of ML models in NLP is vital for managing and interpreting vast amounts of unstructured text data. However, traditional methods often face interoperability, scalability and efficiency challenges, limiting their effectiveness in real-world scenarios (5).

Healthcare is a critical global concern and headaches, particularly migraines, are recognized as one of the most common and disabling conditions worldwide (6,7). The World Health Organization (WHO) reports that nearly half of the global population experiences some form of headache, including tension-type headaches, cluster headaches and migraines (8). Migraines, often episodic, significantly impact individuals’ quality of life and create a substantial economic burden on society (9).

With the rise of social media platforms such as Twitter, individuals increasingly share their personal experiences, including their health struggles (10,11). This presents a unique opportunity to analyze real-time data on conditions like migraines, providing insights into behavioural patterns and emotional responses. Textual data from Twitter can capture individuals’ perceptions of migraine attacks, offering valuable information that can supplement existing medical knowledge and potentially improve predictive capabilities in public health (12).

While existing studies have explored various aspects of migraine epidemiology in the Arab world, there is a need to synthesize these findings, mainly using modern technologies such as SA and ML (13–16). Twitter data, especially in Saudi Arabia, where there is a significant number of active users, provides an opportunity to explore how individuals discuss their migraine experiences (17). However, extracting useful information from unregulated platforms like Twitter poses significant challenges, particularly in filtering relevant tweets and pre-processing Arabic language data (5,18,19). Moreover, current ML models often face difficulties integrating diverse data sources and handling large-scale datasets, limiting their scalability and flexibility (20–22).

Existing SA studies in healthcare often face challenges in effectively analyzing social media data due to the diverse nature of Arabic dialects, the volume of unstructured text and the dynamic nature of social discussions. While approaches such as MARBERT and deep neural networks (DNNs) have shown success in structured datasets, the scalability and interoperability of these models remain limited when applied to real-world, large-scale, unstructured data environments like Twitter (23).

Most frameworks focus solely on individual models, overlooking the potential benefits of multi-agent-based frameworks that coordinate and integrate diverse ML models (24–27). This research bridges the gap by proposing MASHA (Multi-Agent System for Healthcare Sentiment Analysis), a novel multi-agent-based SA system that addresses the volume and complexity of Arabic healthcare-related tweets, specifically targeting migraine-related discussions.

This study presents the following contributions:

1. **Framework Development:** Introduction of **MASHA**, a novel Multi-Agent System (MAS) designed to manage the entire sentiment analysis workflow, from data collection and pre-processing to model training and decision-making.
2. **Scalability and Flexibility:** Integrating multiple ML models (support vector machines (SVM), Naive Bayes (NB) and Logistic Regression (LR)) through ensemble learning within a multi-agent framework to improve sentiment classification in resource-limited healthcare environments.
3. **Case Study Validation:** The proposed framework was applied to real-world Arabic tweets related to migraines, demonstrating improved accuracy, precision, recall and F1-score compared to traditional single-model approaches.
4. **Addressing Dialectal Complexity:** Implement customized pre-processing techniques to handle Arabic dialect variations, enhancing text normalization and feature extraction processes.

MASs involve multiple autonomous agents collaborating to achieve specific goals. In the context of ML, MAS facilitates the integration of diverse models by allowing agents to manage different aspects of the process (28). Prior research has demonstrated the potential of MAS in improving the efficiency and scalability of ML model integration (29–31).

Ensemble learning and hybrid models have traditionally been used to combine the strengths of different ML models, improving overall performance (32–34). However, these methods often struggle with scalability and flexibility in dynamic environments. In NLP applications, ensemble methods face challenges in coordination and efficiency during integration (32).

Despite advancements in ML model integration, existing methods struggle with handling large-scale and dynamic data environments, particularly in NLP applications (5). This study addresses these gaps by proposing a flexible and scalable multi-agent-based framework for integrating diverse ML models.

Recent advancements in SA and ML for healthcare applications have demonstrated the potential of analyzing patient experiences through social media data (35). AlMuhaideb et al. (2023), introduced a novel HoPE-SA dataset that gathers patient experience sentiments from Saudi Arabia’s healthcare services using Arabic Twitter data. The study fine-tuned transformer-based models, such as MARBERT, achieving high accuracy in sentiment classification. This research underscores the growing role of social media SA in understanding patient experiences, particularly in healthcare contexts (36).

Similarly, Khan et al. (2024) applied various ML models, including SVM, decision trees (DT) and DNNs, to classify different types of migraines using patient data. The study employed data augmentation techniques to expand the dataset, significantly improving the model’s accuracy. Notably, the DNN model achieved an accuracy of 99.66%, emphasizing the potential of ML in diagnosing and predicting conditions such as migraines (15). These studies highlight the importance of leveraging advanced ML models and SA to improve the accuracy and scalability of healthcare-related NLP tasks. This aligns with the focus of this research on developing a multi-agent-based framework for SA of Arabic tweets related to migraine detection.

In summary, integrating ML models in NLP is a complex task that requires robust frameworks to manage diverse data and model types effectively (37). This study proposes **MASHA**, a multi-agent-based framework that addresses the challenges of ML models integration. A case study on sentiment classification for migraine detection using Arabic tweets is used to demonstrate the effectiveness of this framework, which enhances both performance and efficiency in integrating diverse models.

The remainder of this study is structured as follows: Section 2 reviews the related work, focusing on previous studies involving MAS, ML integration in NLP and SA, particularly in the context of Arabic language and healthcare data. Section 3 outlines the methodology to develop the multi-agent-based framework for sentiment classification of migraine-related tweets, including data collection, filtering and integrating diverse ML models. Section 4 presents the results of the experiments and discusses the proposed framework’s performance using key metrics such as accuracy, precision, recall and F1-score. Finally, Section 5 concludes the study, summarizing the key contributions and suggesting potential avenues for future research.

## 2. Related Work

The integration of ML models into various applications has been extensively researched, particularly in the domain of NLP. However, there is room for improvement in healthcare-related tasks like SA of social media data for public health monitoring. This section explores the existing MAS, SA and ML model integration work, specifically in NLP and healthcare domains.

### 2.1 Multi-Agent Systems (MAS)

MASs are composed of multiple autonomous agents interacting to achieve common or individual goals (38). In ML contexts, MAS has shown great potential in managing complex processes by distributing tasks among agents. In their study (39), they laid the foundation for MAS by defining key concepts and applications, which have since expanded to various domains such as robotics, distributed computing and decision-making. MAS is particularly advantageous for integrating multiple ML models, as agents can manage different aspects of the integration process, improving system efficiency and scalability.

The key characteristics of agents in MAS, autonomy, reactivity, pro-activeness and social ability, enable them to operate independently, respond to changes, take initiative and collaborate effectively (28). These traits are essential for efficiently integrating ML models in the MASHA framework for SA and healthcare applications. Figure 1 visualizes these core characteristics, illustrating how agents’ attributes support effective collaboration and task execution within the system.

**Figure 1.**
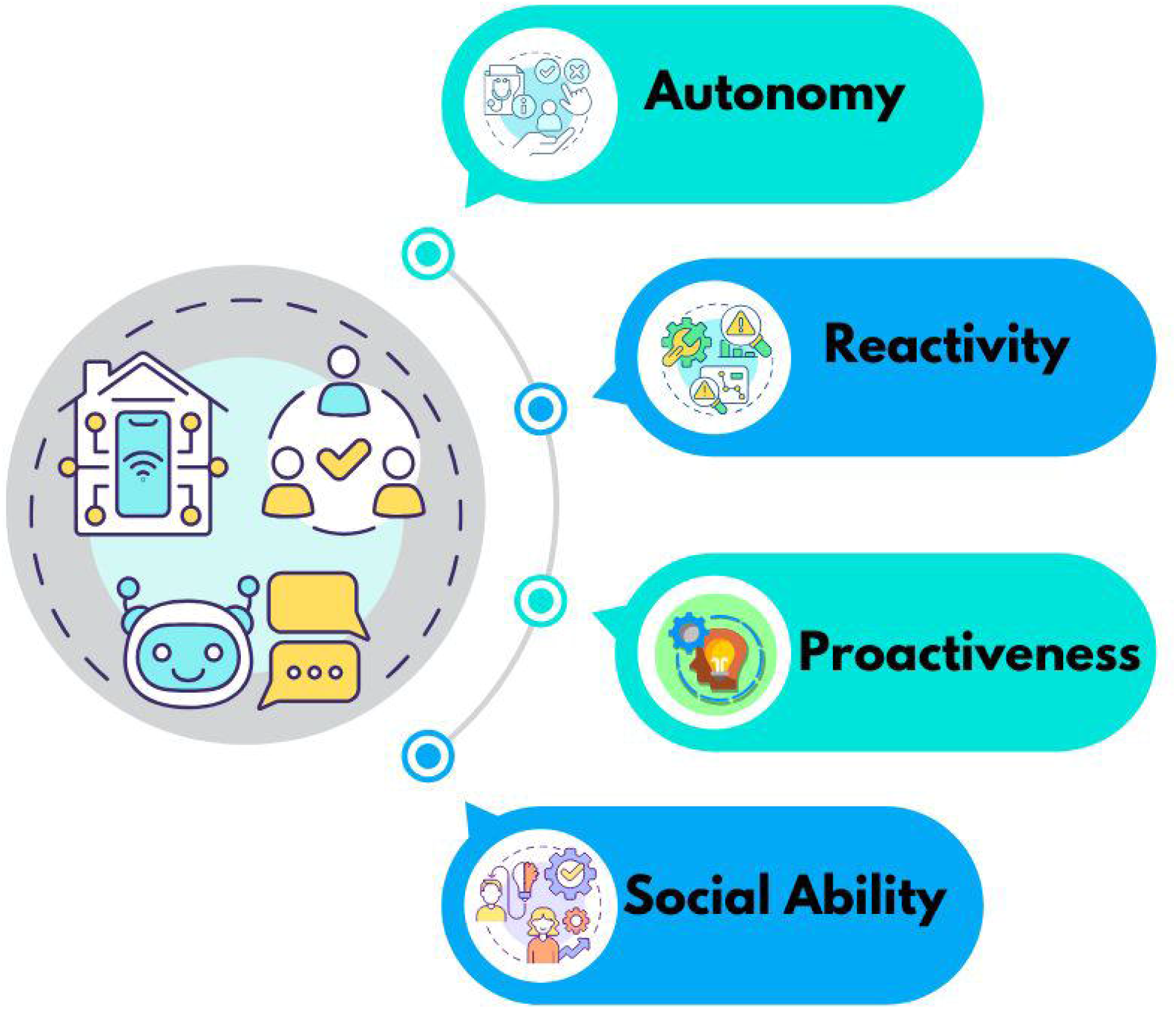
Core Characteristics of Agents in MAS

In recent years, MAS has been employed to address challenges in data processing and integration of models in real-time systems (29). For example, MAS has been used in medical applications to diagnose and process patient data, by allowing different agents to handle diverse tasks such as data pre-processing, model development and model integration, MAS can significantly reduce bottlenecks and improve performance, especially when managing large datasets or complex data streams (41). Given these strengths, applying MAS in NLP tasks such as SA can significantly enhance system flexibility and scalability.

### 2.2 Sentiment Analysis (SA)

Sentiment Analysis (SA) is a popular approach in NLP that classifies text based on individuals’ emotions, opinions or sentiments. SA has been widely applied across industries to gauge consumer feedback, track public opinion and monitor trends. In healthcare, SA can be particularly valuable for monitoring patient experiences and disease-related discussions on social media platforms (42).

Arabic sentiment analysis (ASA), in particular, presents unique challenges due to the language’s rich morphological structure and the prevalence of dialects in social media conversations (43). Early research on SA in Arabic faced significant difficulties due to the lack of resources such as sentiment lexicons and annotated corpora (44). Also (45), provided a comprehensive overview of SA techniques while (46), discussed the importance of combining ML with NLP to enhance the accuracy of Arabic text classification.

Recent studies have explored deep learning (DL) techniques for ASA (47). The authors (48) surveyed the latest advances in this field, noting an increasing trend toward DNN for feature extraction and sentiment classification. However, these studies often focus on general-purpose applications rather than domain-specific tasks like healthcare. This gap highlights the need for further research on applying SA techniques to specific problems like migraine detection.

### 2.3 ML Integration in NLP

Extensive research has been done on integrating multiple ML models, especially in ensemble learning techniques like bagging, boosting and stacking (32,49). These methods combine different models’ strengths to enhance prediction accuracy and robustness. However, despite their success, traditional ensemble methods face challenges when applied to large-scale and dynamic environments, such as social media data in healthcare contexts (50).

In NLP, ensemble learning methods have been used to combine the predictive power of different algorithms, particularly for tasks like SA, text classification and machine translation (51,52). However, these approaches often have inefficiencies in coordinating multiple models, particularly when handling heterogeneous data types and formats (53). The complexity of coordinating ML models increases when dealing with healthcare-related data, as the accuracy and timeliness of insights are crucial for improving patient outcomes (35).

Several studies have proposed hybrid models that combine ML with rule-based systems to handle complex NLP tasks better (54). However, these models often lack the flexibility and scalability to process real-time data or handle dynamic input like social media streams. As a result, there is a growing interest in using MASs to manage the integration of ML models in such tasks. By distributing the workload across different agents, MAS can help address coordination, scalability and real-time processing issues.

### 2.4 Gaps in Current Research

Despite advances in SA, ML integration and MAS, current research faces several limitations when applied to healthcare-related NLP tasks like migraine detection. Traditional ML model integration methods often struggle with the sheer volume and variety of social media data, especially when processing real-time inputs from platforms like Twitter (42). Moreover, there is a shortage of high-quality Arabic datasets for NLP tasks, further complicating the development of effective sentiment classification models for Arabic-speaking populations (46).

Current SA models, particularly in Arabic, still face challenges in handling dialect variations, the lack of standardized lexicons and the complexity of Arabic morphology (44). Additionally, existing multi-agent frameworks, while promising, have yet to be fully exploited for healthcare-related SA. Specifically, their application in NLP tasks like migraine detection using Arabic tweets remains largely unexplored.

These gaps underscore the need for more flexible, scalable and context-aware frameworks that integrate diverse ML models to handle real-time healthcare data in dynamic environments. This research proposes a multi-agent-based framework to address these challenges, leveraging the strengths of MAS for efficient ML model integration in NLP tasks related to public health monitoring.

## 3. Methodology

This section describes the proposed **MASHA**, a multi-agent-based framework for integrating ML models in the context of SA for migraine detection using Arabic tweets. **MASHA** is designed to be scalable, flexible and efficient, leveraging multiple agents to handle different aspects of data acquisition, pre-processing, model development and decision-making. The architecture consists of three key layers: the Data Pre-processing Agents Layer, the Model Development Agents Layer, and the Decision-Making Agents Layer. Each agent in the **MASHA** framework is responsible for specific tasks to ensure the overall efficiency and performance of the system, enabling seamless integration of ML models and effective SA in real-time.

### 3.1 Overview of the Multi-Agent Framework

The proposed multi-agent framework consists of several autonomous agents, each responsible for specific tasks in the sentiment classification process. Figure 2 illustrates the overview of the **MASHA** framework architecture.

**Figure 2.**
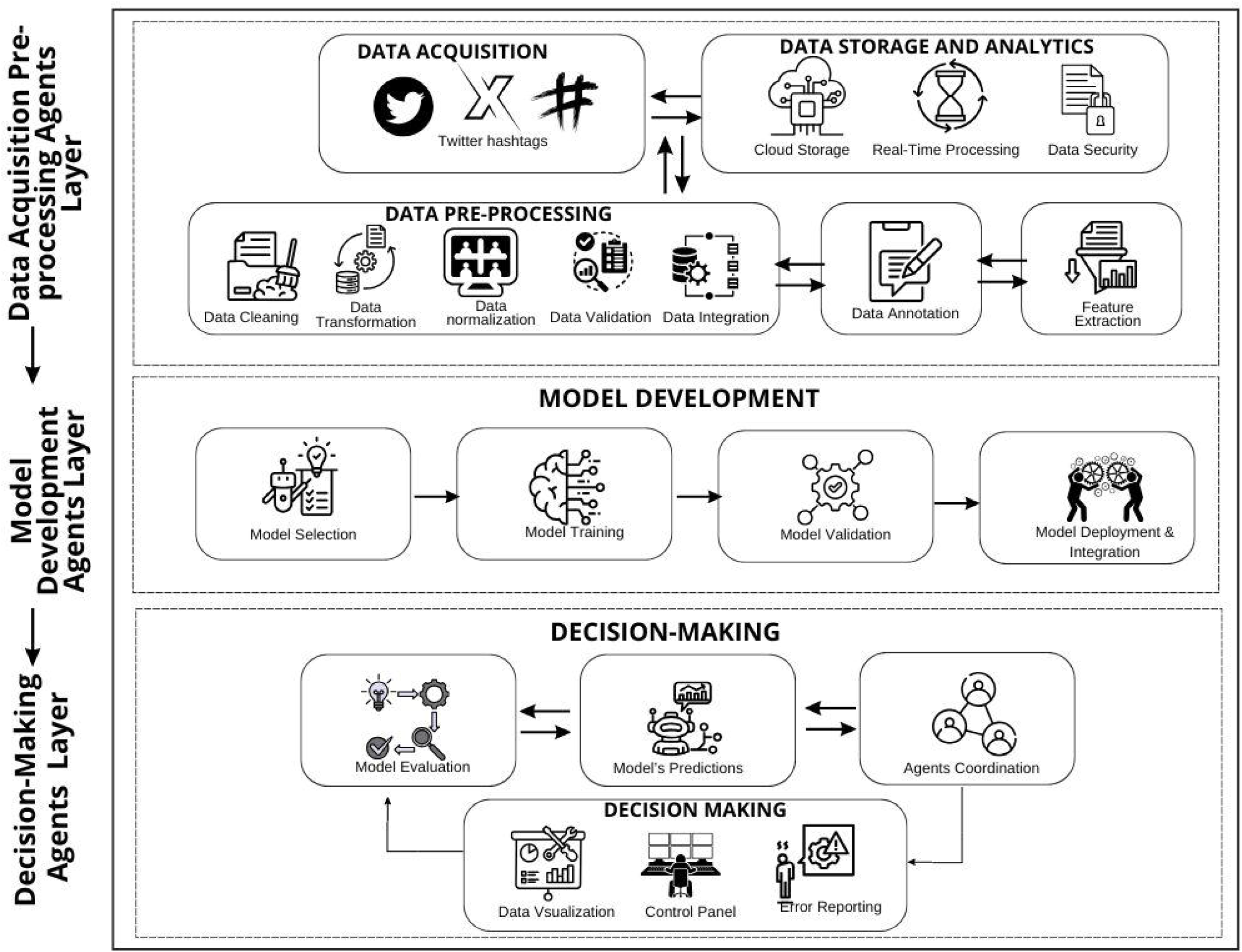
Overview of the MASHA Framework Architecture

#### 3.1.1. Data Acquisition and Pre-processing Agents Layer

The data pre-processing layer handles all tasks related to collecting and preparing data for further analysis. This includes the following components:

- **Data Acquisition Agent**: This agent is responsible for gathering relevant tweets from Twitter. It tracks specific hashtags related to migraine and healthcare, ensuring the dataset contains real-time and historical tweets from Arabic-speaking users. The agent interacts with the Twitter API to continuously update the dataset. Figure 3 displays a word cloud generated from the dataset, highlighting the most frequently mentioned terms related to migraines.
- **Data Storage and Analytics**: After data is acquired, it is stored in a cloud-based system that supports real-time processing and ensures data security. Cloud storage enables the system to scale dynamically as more data is collected. The data analytics module prepares the raw data for the pre-processing pipeline.
- **Data Pre-processing Agent**: This agent performs essential tasks such as:
  - **Data Cleaning**: Removing irrelevant information such as URLs, mentions, non-Arabic characters and special symbols.
  - **Data Transformation**: Converting data into a consistent format that subsequent modules can process.
  - **Data Normalization**: Standardizing the text by handling variations in spelling, repeated letters and dialect-specific expressions in Arabic.
  - **Data Validation and Integration**: Ensuring the collected data is relevant and accurate and then integrating it into the system for further processing.
  - **Data Annotation**: Human annotators label the data, classifying tweets as positive or negative. This creates a gold standard dataset for model development.
  - **Feature Extraction**: The system applies Term Frequency-Inverse Document Frequency (TF-IDF) vectorization and word embeddings to convert raw text into feature vectors suitable for ML models. Specifically, we applied AraVec, a pre-trained Arabic word embedding model, to generate dense vector representations of words. These embeddings capture semantic similarities and contextual relationships between words, enhancing the sentiment classification accuracy for Arabic tweets.

**Figure 3.**
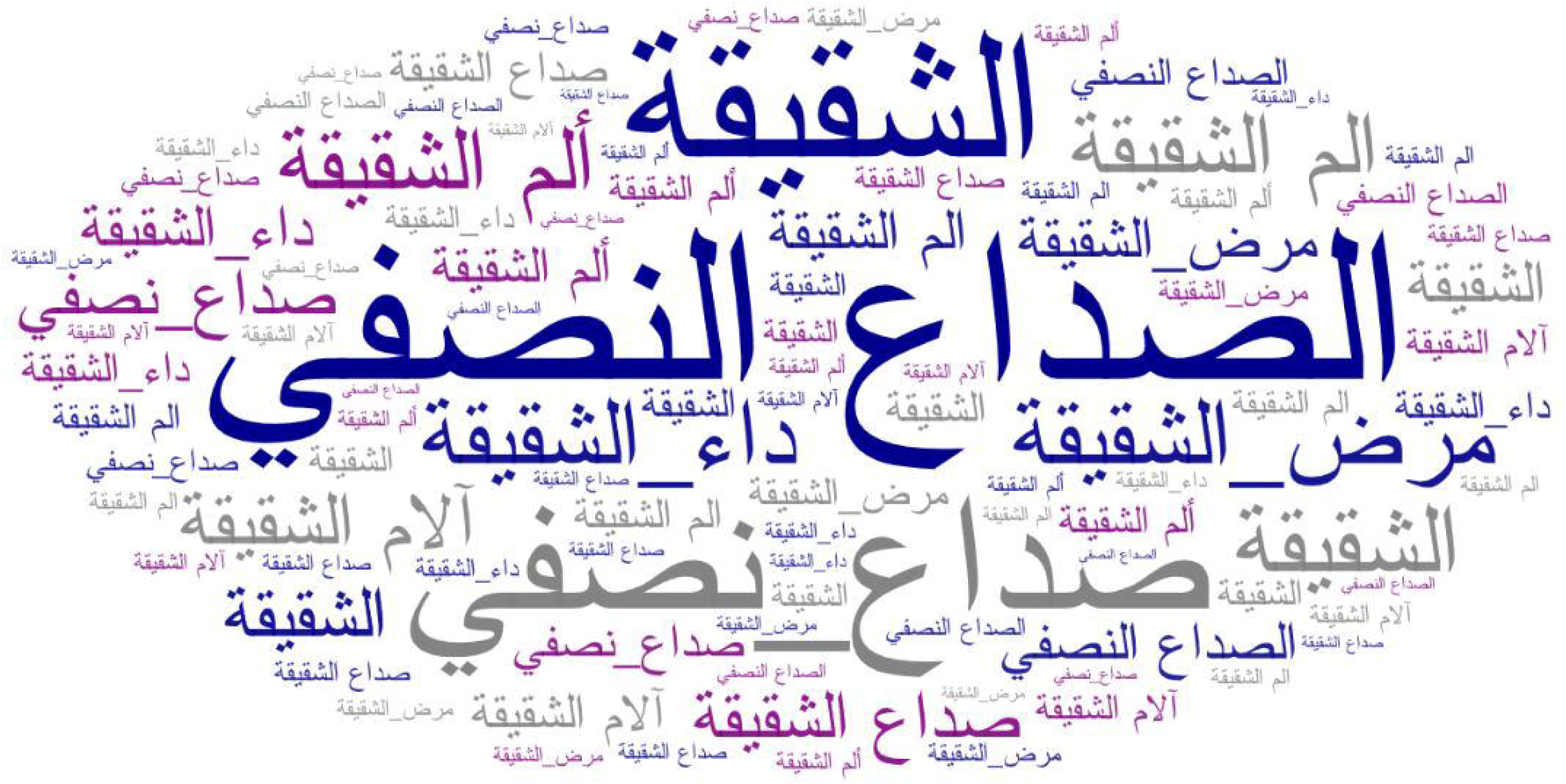
Word Cloud of Migraine-Related Terms in Arabic Tweets. The word cloud displays Arabic terms related to migraines. The most prominent terms include: الصداعالنصفي (migraine), الصداعالنصفي (migraine headache), ألمالشقيقة (migraine pain), داءالشقيقة (migraine disease), ? صداعنصفي (half-sided headache) and مرضالشقيقة (migraine illness).

#### 3.1.2 Model Development Agents Layer

ML models are selected, trained, validated and deployed in the model development layer. The components of this layer include:

- **Model Selection Agent**: This agent chooses appropriate ML algorithms for the sentiment classification task. The framework employs several ML models, including SVM, NB and LR. The selection is based on the nature of the data and the classification task. The algorithms used in the framework are as follows:
  - **SVM:** obtains an optimal hyperplane that separates classes with maximum margin. The main steps in SVM are:
    - Calculate the decision boundary by solving the optimization problem.
    - Apply the kernel trick (linear, polynomial or RBF) for non-linear data if required.
    - Classify the sentiment based on the hyperplane’s side.
  - **NB**: is based on Bayes’ theorem assuming independence among features. It calculates the probability of each sentiment class ***C*** given the features ***X***:

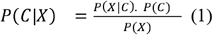

The class with the highest probability is chosen.
  - **LR:** is used to estimate probabilities using the sigmoid function:

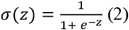

Where *z* = *w*^*T*^*X+ b* . The model predicts positive or negative sentiment based on whether σ(*z*) is above a threshold (usually 0.5).

- **Model Development Agent**: After selecting the models, the training agent uses the annotated and pre-processed data to train each model. The training process includes splitting the dataset into training and testing sets to evaluate the model’s performance and avoid overfitting.
- **Model Validation Agent**: The trained models are validated against a separate validation set. The validation agent assesses each model’s accuracy, precision, recall and F1-score to determine its effectiveness in classifying sentiment from migraine-related tweets. The formulas used for these metrics are as follows:
  - **Accuracy**:

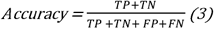

where *TP* is True Positives, *TN* is True Negatives, *FP* is False Positives and *FN* is False Negatives.
  - **Precision:**

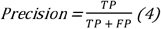
  - **Recall:**

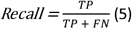
  - **F1-score:**

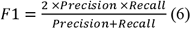
  - **Model Deployment and Integration Agent**: Once models are validated, they are deployed into the system. This agent handles the integration of multiple models through ensemble techniques such as voting or stacking, improving overall classification accuracy by combining the strengths of each model.

#### 3.1.3 Decision-Making Agents Layer

The decision-making layer coordinates the predictions and outputs generated by the model development layer, allowing the system to deliver actionable insights based on the SA of tweets. It consists of the following components:

- **Model Evaluation Agent**: This agent is responsible for continuously evaluating the integrated model’s performance. It monitors the ensemble model’s predictions and continuously calculates performance metrics such as accuracy, precision, recall and F1-score to ensure the system operates optimally.
- **Decision-Making Agent**: This agent oversees the overall decision-making process based on the evaluated predictions. It coordinates the activities of all other agents and generates final predictions visualized on a control panel for stakeholders. The system also allows for error reporting, making it possible to identify and address issues in real-time.
- **Data Visualization and Reporting**: This module visualizes the predictions and trends identified by the ML models. It enables stakeholders to gain insights into the general sentiment about migraines and provides a control panel for monitoring system performance. The reporting module offers transparency into decisions and provides error reporting to alert administrators of any issues.

### 3.2 Dataset

#### 3.2.1 Data Collection

The dataset used in this study was collected from Twitter, focusing on Arabic tweets related to migraines. The Twitter API was utilized to retrieve tweets in real-time and historical data based on predefined hashtags and keywords. The primary hashtags used for data collection included # صداع نصفي (#migraine), # صداع (#headache), # صداع الشقيقة (#migraine) and other health-related terms frequently associated with migraine discussions. The tweets were collected from March to May 2021, and only tweets written in Arabic were included in the dataset.

To ensure relevance, tweets were filtered based on geo-tags and user profile information to select content related to Saudi Arabia. The ease of use, availability and accuracy of the Twitter API made it ideal choice for data collection. We applied for a developer account to access the API and created an application to obtain the necessary OAuth parameters: “consumer key”, “consumer secret”, “access token” and “access secret”.

Python programming languages, related software packages and Microsoft Excel were used to collect tweets, preprocess them and build the annotated dataset. Verified users and accounts with significant activity were prioritized to filter irrelevant content, improving the quality of the collected data.

#### 3.2.2 Data Characteristics

The dataset consisted of approximately 14,000 Arabic tweets. Each tweet was accompanied by metadata, including the tweet’s timestamp, user information, location and the specific hashtag or keyword used. The collected data included real-time and historical tweets, providing a comprehensive view of how users discuss migraines over time. The geographic focus was on users from Saudi Arabia, where Twitter usage is exceptionally high and healthcare discussions are active. Figure 4 provides a snapshot of this study’s sentiment classification dataset. The dataset consists of Arabic tweets related to migraines, annotated for sentiment as either positive or negative. This sample highlights the variety of textual data processed, showcasing the different structures and expressions found in the collected tweets, including the dialectical variations that pose a challenge for classification models.

**Figure 4.**
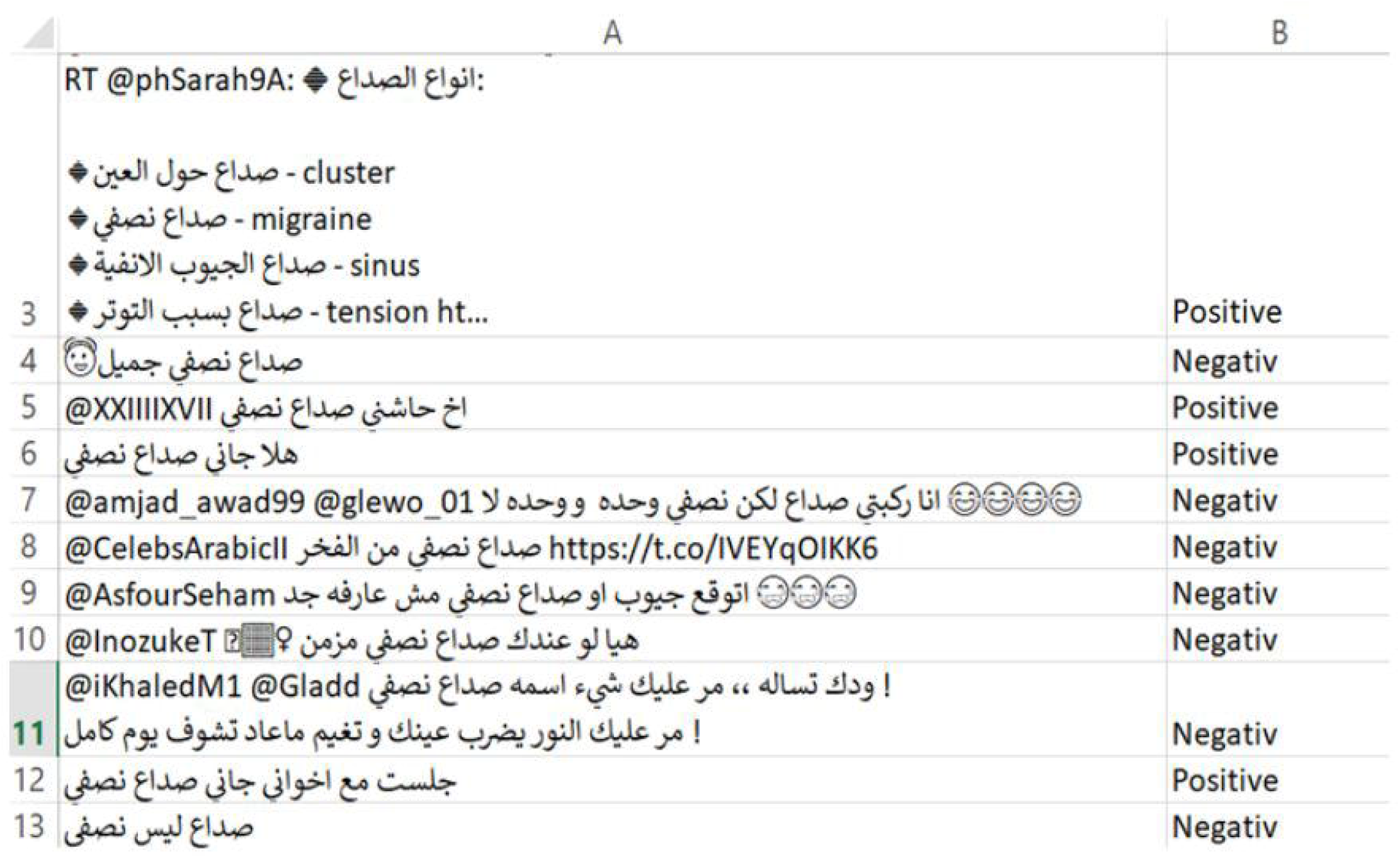
Sample of the dataset

The dataset sample in Figure 4 shows Arabic tweets with their sentiment labels. English translations are provided below:

□ Row 3: “RT @phSarah9A: ⍰ Types of headaches: ⍰ cluster headache - cluster ⍰ half- sided headache - migraine ⍰ frontal sinus headache - sinus ⍰tension headache caused by stress - tension ht…” - (Positive)
□ Row 4: “Half-sided headache is beautiful ⍰” - (Negative)
□ Row 5: “I have half-sided migraine headache @XXIIIIXVII” - (Positive)
□ Row 6: “I’m cured, thank God, of half-sided headache” - (Positive)
□ Row 7: “I’m really suffering from half-sided headache but alone and alone and alone lol ⍰ ⍰ ⍰ ⍰@amjad_awad99 @glewo_01” - (Negative)
□ Row 8: “@CelebsArabicII Half-sided headache from envy https://t.co/IVEYqOlKK6” - (Negative)
□ Row 9: “@AsfourSeham Expect pockets or half-sided headache even if he knows you ⍰ ⍰ ⍰” - (Negative)
□ Row 10: “@InozukeT ⍰ ⍰ ♀□? You or anyone else have chronic half-sided headache” - (Negative)
□ Row 11: “@iKhaledM1 @Gladd Half-sided headache, what’s its name, oh my God you’re on my mind! I wish you all the best, may your eyes see the light of a perfect day” - (Negative)
□ Row 12: “I sat with my brothers with half-sided headache” - (Positive)
□ Row 13: “It’s a headache, not half-sided” - (Negative)

#### 3.2.3 Pre-processing

Given the unstructured nature of Twitter data and the linguistic complexities of Arabic, extensive preprocessing was performed on the collected dataset. The following pre-processing steps were applied:

- **Text Normalization**: The normalization process involved removing diacritics, handling repeated characters and standardizing the spelling of common words related to migraines. Variants such as “الشقيقة” (migraine) and “شقيييقة” (miiigraine - with repeated letters) were normalized to a single standard form to enhance consistency and model accuracy. Furthermore, mentions, unrelated hashtag, punctuation, URLs and non-Arabic words were removed to reduce noise. However, relevant hashtags (e.g., # صداع_نصفي(#migraine_headache)) were retained to capture crucial context related to migraines.
- **Data Validation and Integration:** Involves ensuring that the collected data is relevant, accurate and free from duplicates. Validation checks include confirming that tweets contain relevant migraine-related content by verifying specific keywords and hashtags. Duplicate tweets, bot-generated spam and unrelated texts are filtered out during this process. Integration involves merging real-time and historical data streams into a unified dataset, ensuring consistency in formats such as timestamps, text encoding and metadata fields. This step ensures that the final dataset is cohesive and ready for subsequent pre-processing and analysis.
- **Tokenization**: Each tweet was broken down into individual tokens (words) to allow for feature extraction.
- **Stemming and Lemmatization**: The “Arabic Stanford Lemmatizer” was used to standardize the text by reducing words to their base forms. This helped address the morphological complexity of the Arabic language, which is essential for handling various forms of the same word. Moreover, custom rules were applied to handle common dialectal variations and spelling inconsistencies. This confirmed that feature extraction was more consistent, leading to improved classification accuracy.
- **Stop-Word Removal**: Common Arabic stop words that do not contribute to the sentiment (e.g., prepositions, conjunctions) were removed to focus on the meaningful content of the tweets.

Below is an example of how a tweet from the dataset undergoes these steps:

### Original tweet

*“عندي صداع نصفي شديد جدااااا والله #الشقيقةhttps://migraineinfo.com“*.

English translation: *“I have a very severe migraine, I swear to God* Ill *#migraine https://migraineinfo.com“*.

1. **Data Cleaning:** The tweet is cleaned by removing unwanted elements, such as URLs, hashtags and emojis: *“* عندي صداع نصفي شديد جدا والله*”*. English translation: *“I have very severe migraine I swear to God”*.
2. **Text Normalization:** Spelling variations and repeated letters are standardized: *“* عندي صداع نصفي شديد جدا والله*”*. English translation: *“I have very severe migraine I swear to God”*.
3. **Feature Extraction:** The cleaned and normalized text is transformed into feature vectors using TF-IDF or word embeddings. For example: *[0*.*14, 0*.*08, 0*.*30,]*

This example demonstrates how the preprocessing steps convert unstructured text into a structured input format that the ML models can process efficiently.

#### 3.2.4 Class Distribution and Labelling

A group of annotators manually annotated each tweet’s sentiment. The tweets were classified into two sentiment categories: “Positive” and “Negative”. The manual annotation process ensured that each tweet was accurately labelled based on its sentiment toward migraine experiences. The final distribution of the dataset is shown in Figure 5:

**Figure 5.**
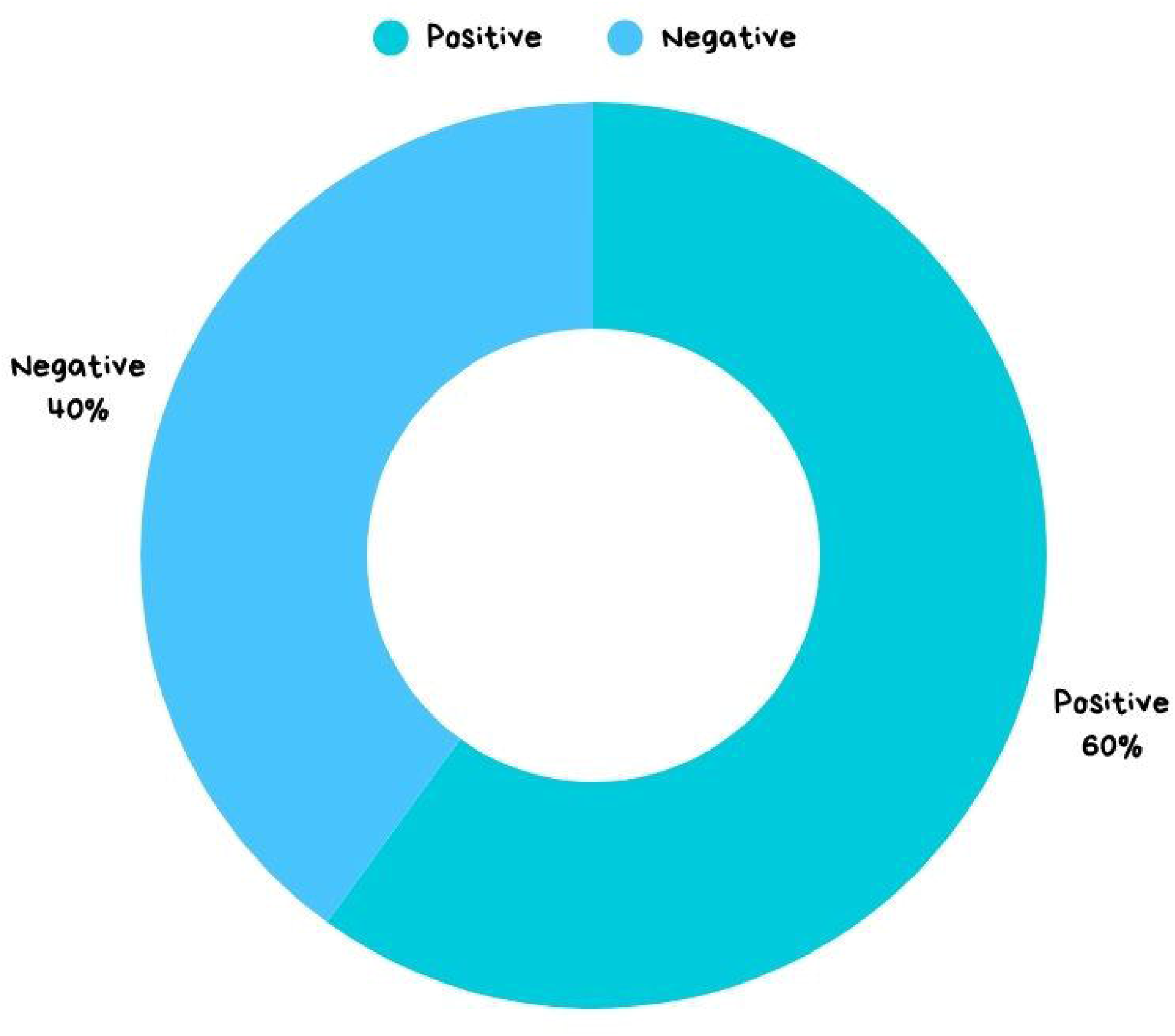
Sentiment Distribution in Arabic Tweets

The classification was based on specific keywords, context, and sentiment-related expressions. Table 1 presents the features and symptoms that were used to distinguish between “Positive” and “Negative” tweets:

**Table 1.** Classification Features for Positive and Negative Tweets.

| Positive | Negative |
| --- | --- |
| Contains the word “صداع نصفي” (migraine) indicating the user is experiencing or has experienced a migraine. | Contains the word “صداع نصفي” (migraine) but used in a general context without indicating a personal experience (e.g., informational tweets). |
| Includes words or phrases indicating symptoms | Includes words that describe unrelated contexts |
| such as “الم” (pain) or “صداع قوي” (strong headache). | (e.g., “My friend has migraine” or “تعرفوا على أعراض الصداع النصفي” – (Learn about migraine symptoms)). |
| Emojis or hashtags that imply distress (e.g., 😞, 🤯). | Neutral or unrelated hashtags (e.g., #صحة(#health), #معلومات (information)). |

The classification process aimed to accurately reflect user sentiment and ensure the dataset contained relevant examples for training and testing the SA models.

### 3.3 Challenges and Limitations

Several challenges were encountered during the data collection and preprocessing stages. The primary challenge was the linguistic complexity of Arabic, including dialectal variations, which required extensive normalization and stemming. Additionally, Twitter’s unregulated nature made it difficult to filter irrelevant or off-topic tweets. However, the MAS’s preprocessing agent efficiently handled these issues, ensuring the final dataset was clean and relevant for SA.

## 4. Results and Discussion

This section presents the results of the proposed multi-agent-based framework for SA on migraine-related Arabic tweets. It evaluates the framework’s performance in key metrics such as accuracy, precision, recall and F1 score. The results are analyzed in comparison with traditional methods, emphasizing the improvements brought by the MAS. Additionally, this section discusses the implications of the findings, particularly in the context of healthcare and public health.

### 4.1 Sentiment Classification Results

The dataset was divided into 70% for training, 15% for validation and 15% for testing sets to ensure balanced evaluation. The training set was used to fit the models, while the validation set was used for hyperparameter tuning and preventing overfitting. The testing set was reserved for final model performance evaluation.

The training process involved splitting the data into batches, with a batch size of 32 for SVM, NB and LR models. The models were trained for (50) epochs with early stopping applied to prevent overfitting. The learning rate was set at (0.001) and the Adam optimizer was used for gradient updates. Furthermore, 5-fold cross-validation was used to further assess model robustness.

The sentiment classification results obtained from the models and their integration through ensemble learning techniques are summarized in Table 2. As shown in Figure 6, the ensemble model demonstrated the highest performance across all metrics, achieving an accuracy of 90.0% and an F1-score of 89.46%. The ensemble approach improved overall classification performance by effectively balancing precision and recall, as reflected by the increase in both metrics across all models.

**Table 2.**
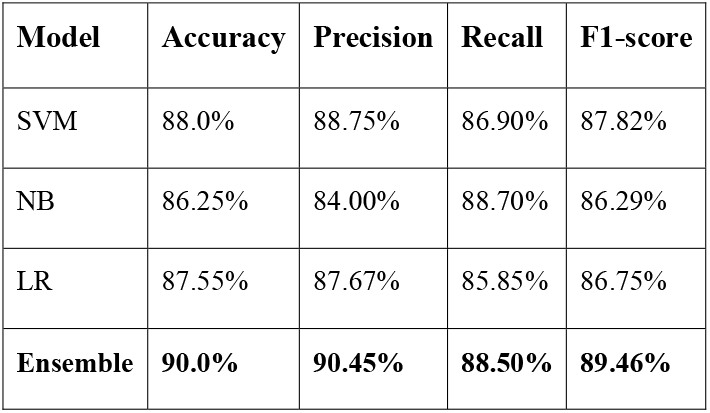
Performance of Individual Models and the Ensemble Model.

| Model | Accuracy | Precision | Recall | F1-score |
| --- | --- | --- | --- | --- |
| SVM | 88.0% | 88.75% | 86.90% | 87.82% |
| NB | 86.25% | 84.00% | 88.70% | 86.29% |
| LR | 87.55% | 87.67% | 85.85% | 86.75% |
| **Ensemble** | **90.0%** | **90.45%** | **88.50%** | **89.46%** |

**Figure 6.**
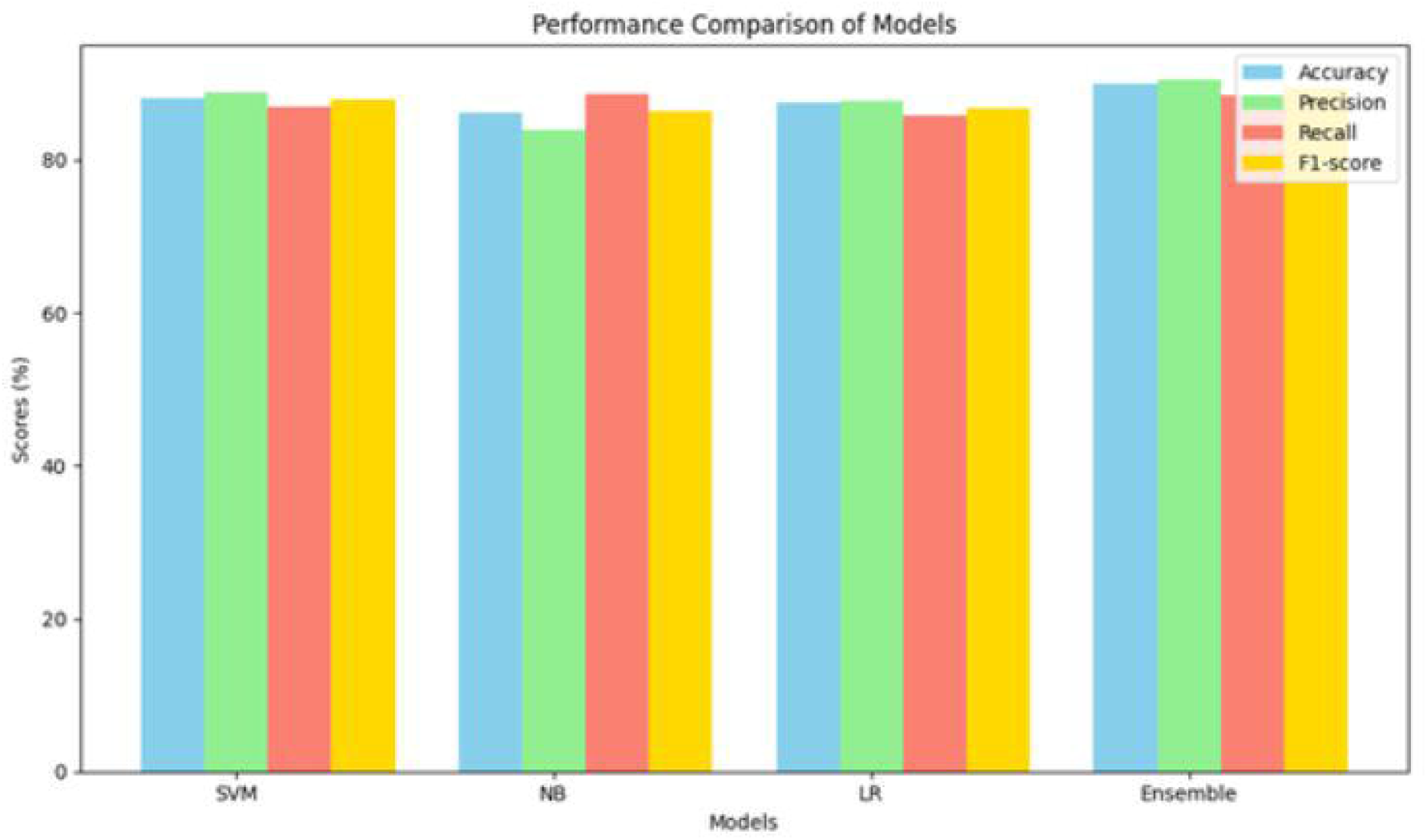
Performance of SVM, NB, LR and Ensemble Models

The ensemble model, which combines the predictions of SVM, NB and LR, achieved the highest performance across all metrics, confirming the effectiveness of model integration through the **MASHA** framework.

In addition to the key performance metrics discussed in Table 1, the confusion matrix provides a more detailed breakdown of the model’s performance in sentiment classification for migraine-related Arabic tweets. As shown in Figure 7, the matrix displays the correct and incorrect classifications made by the model, with 160 correctly classified positive tweets and 110 correctly classified negative tweets. However, some misclassifications occurred, with 21 negative tweets misclassified as positive and 24 positive tweets misclassified as negative.

**Figure 7.**
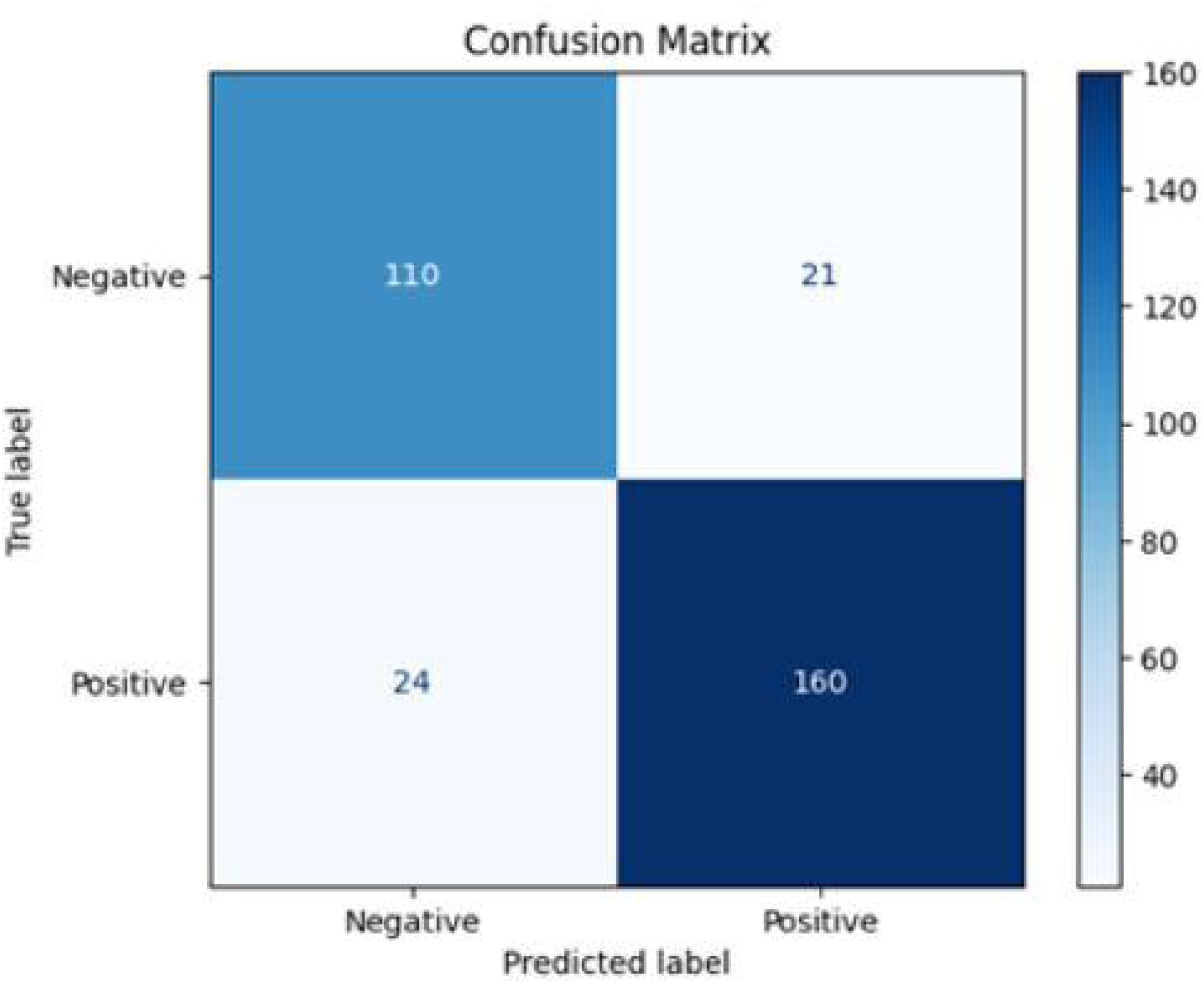
Confusion Matrix for Sentiment Classification on Test Set (300 tweets)

### 4.2 Comparison with Traditional Approaches

Compared to traditional sentiment classification methods that typically rely on a single machine learning model, the multi-agent-based framework, **MASHA**, offers several key advantages:

- **Scalability**: The proposed system can handle large volumes of tweets in real-time thanks to the division of tasks among multiple agents. Each agent specializes in a specific task, allowing efficient processing without overwhelming a single system component.
- **Flexibility**: By integrating multiple ML models, the system is more adaptable to changes in data and can accommodate the diverse linguistic variations in Arabic, including dialects commonly used in Saudi Arabia.
- **Improved Accuracy**: The ensemble learning technique outperformed traditional methods, improving classification accuracy by approximately 2.5%. This improvement is significant in healthcare applications, where accuracy is crucial for making reliable predictions.

### 4.3 Error Analysis

Despite the MASHA framework’s overall solid performance, particular challenges impacted classification accuracy. As depicted in Figure 8, the primary sources of error included handling Arabic dialect variations (40% of errors) and the short length of tweets (30% of errors), making it difficult to accurately assess sentiment.

**Figure 8.**
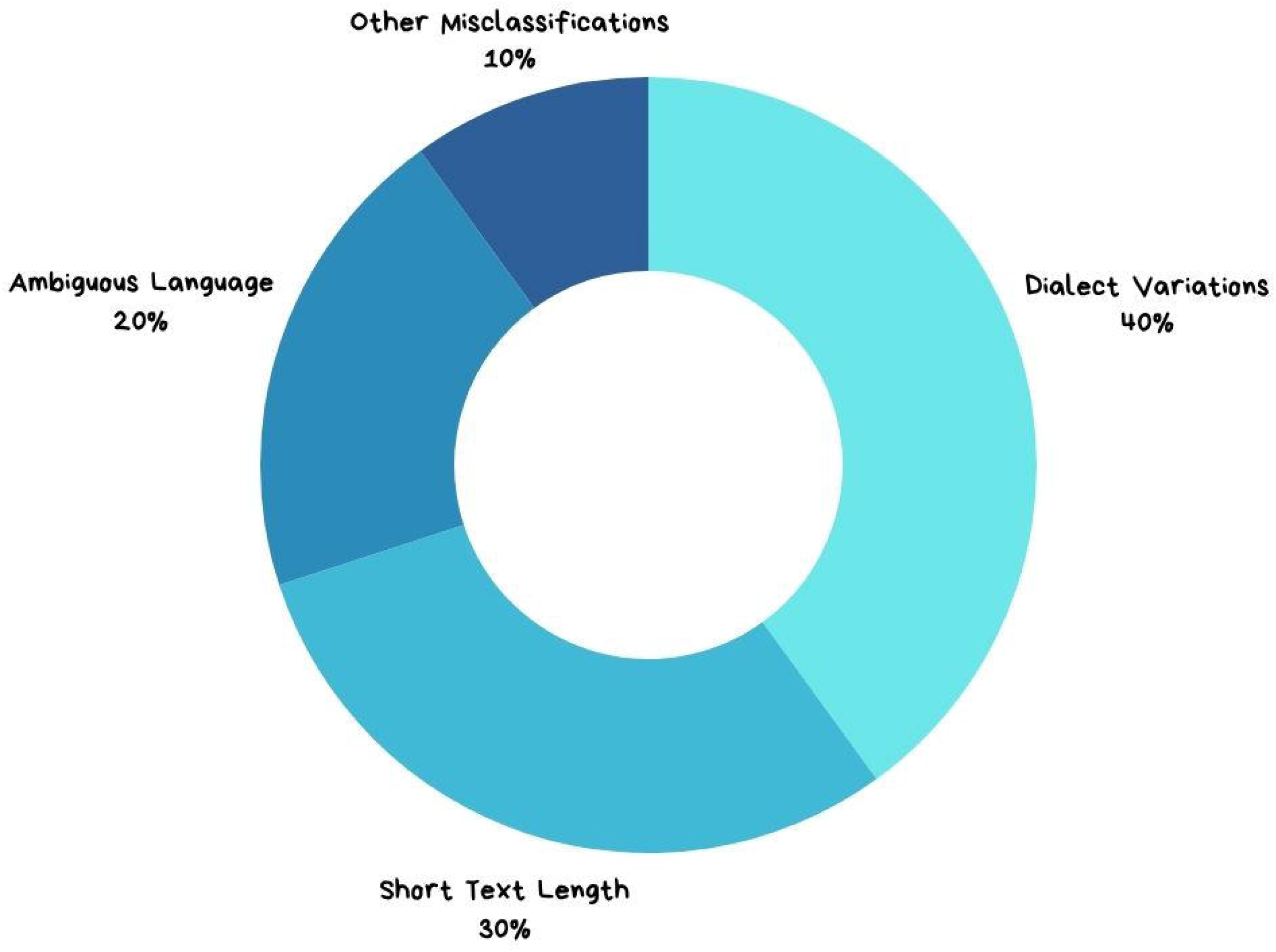
Error Distribution in Sentiment Classification

- **Handling Arabic Dialects**: Dialect variation is one of the primary challenges in SA for Arabic tweets. Although the pre-processing agent performed text normalization and stemming, some dialect-specific expressions were complex to standardize, which impacted the accuracy of sentiment classification in some instances.
- **Short Text Length**: Tweets often contain very short messages, which limits the amount of context available for sentiment classification. As a result, some tweets were misclassified, mainly when they contained ambiguous language or lacked clear sentiment indicators.

During the analysis, several common error types were observed:

- **Ambiguity:** Tweets such as “الحمد لله بعد الصداع ارتحت” (“Thank God I feel better after the headache”) could be interpreted as both positive (relief) and negative (due to mentioning a headache).
- **Sarcasm:** Sentences like “ما في أحلى من صداع نصفي للصباح” (“Nothing better than a migraine in the morning”) were often misclassified due to the sarcastic tone.
- **Incomplete Context:** Some tweets lacked sufficient context for accurate classification, such as “أوه لا … الصداع بدأ ! …’:l O??” (“Oh no… the headache started!”).

Although the dataset primarily contained tweets from “Saudi Arabia”, dialect variations such as “Najdi”, “Hejazi” and “Gulf Arabic” were present. For example, the word “راس” (head) can appear as “راس” (head) in certain dialects due to phonetic spelling. Preprocessing included normalizing these variations to a standard form and the ML models were fine-tuned to recognize common synonyms and spelling differences to minimize misclassification due to dialect-specific terms.

### 4.4 Implications for Healthcare and Public Health

The findings from this study have significant implications for the healthcare sector, particularly in the context of public health monitoring. By analyzing real-time social media data, healthcare providers and policymakers can gain valuable insights into patient experiences with migraines, track public sentiment regarding healthcare services and detect emerging health trends.

- **Improved Patient Monitoring**: SA of social media data allows healthcare providers to monitor how patients perceive their migraine symptoms, treatments and overall healthcare experiences. This can help identify common challenges faced by patients and lead to improvements in treatment strategies.
- **Real-Time Public Health Insights**: The system’s real-time processing of tweets enables public health agencies to identify trends and patterns related to migraine episodes and their impact on society. This can inform public health campaigns and interventions to mitigate migraines’ impact on quality of life.
- **Scalability to Other Healthcare Conditions**: While this study focuses on migraines, the multi-agent-based framework can be adapted to analyze other health conditions, such as influenza, diabetes or mental health issues. The system’s flexibility and scalability make it a valuable tool for broader public health applications.

### 4.5 Limitations and Future Work

Despite the positive results, this study has several limitations that should be addressed in future research:

- **Dataset Size**: Although the dataset used in this study is significant, expanding the dataset to include more diverse sources of Arabic social media data would improve the generalizability of the results. Including data from other platforms like Facebook or Instagram could provide a more comprehensive understanding of public sentiment.
- **Further Model Optimization**: Future work should optimize the ML models used in the framework, particularly in handling complex Arabic language structures and dialects. Exploring DL techniques such as recurrent neural networks (RNNs) or transformers may improve classification performance.
- **Sentiment Lexicon Development**: Given the challenges associated with Arabic dialects, developing a more comprehensive sentiment lexicon specific to Arabic social media would improve sentiment classification accuracy.

**MASHA’s** proposed multi-agent-based framework demonstrated its effectiveness in integrating multiple ML models for SA in healthcare-related tasks. The framework improved classification accuracy and provided valuable insights into patient experiences and public health trends. The system’s scalability and flexibility suggest that it can be expanded for other health-related applications.

### 4.6 Discussion

This study’s results demonstrate the **MASHA** framework’s effectiveness in integrating multiple ML models for SA in healthcare-related tasks. By leveraging ensemble learning techniques, the system achieved higher accuracy, precision and F1-scores than individual models. The key takeaway from the research is the importance of integrating multiple models to balance different performance metrics to achieve an overall better sentiment classification outcome.

- **Healthcare Insights**: The system provides significant insights for the healthcare sector, especially in real-time tracking of patient experiences and public health trends. Analyzing sizeable social media data enables healthcare providers to stay updated on patient sentiment and emerging health conditions.
- **Adaptability**: The modular, adaptable structure of the **MASHA** framework ensures that it can be extended to various healthcare conditions, making it a valuable tool for public health campaigns and interventions. Furthermore, the flexibility in handling multiple Arabic dialects expand the system’s application to different Arabic-speaking regions, further improving its utility for healthcare and public policy.

Addressing the current limitations, such as dataset expansion, further model optimization and developing an Arabic-specific sentiment lexicon, would improve the framework’s performance even further. Additionally, exploring the potential of DL techniques could yield significant gains in handling the complexity of Arabic texts, ensuring better results for SA tasks.

## 5. Conclusion

This paper introduced **MASHA** (Multi-Agent System for Healthcare Sentiment Analysis), a scalable and flexible multi-agent-based framework that integrates multiple ML models for SA in healthcare. The proposed framework was validated through a case study focusing on sentiment classification for migraine detection using Arabic tweets. By leveraging the strengths of multiple ML models, including SVM, NB and LR and employing ensemble learning techniques, **MASHA** demonstrated significant improvements in classification performance compared to traditional methods.

The results showed that the ensemble approach improved the overall accuracy, precision, recall and F1-score, with the ensemble model achieving an F1-score of 89.46 %. This underscores the importance of integrating diverse ML models in handling complex healthcare data, especially when analyzing unstructured text from social media platforms like Twitter. Moreover, using a multi-agent architecture allowed the system to efficiently manage data acquisition, pre-processing, model development and decision-making tasks, ensuring the system’s scalability for large-scale, real-time SA.

The implications of this study extend beyond migraine detection. **MASHA’s** adaptability makes it suitable for analyzing sentiment in various healthcare conditions, providing valuable insights for healthcare providers and policymakers. By analyzing real-time social media data, **MASHA** offers a powerful tool for tracking public sentiment, improving patient monitoring and informing public health decisions.

However, this research also highlighted several challenges, particularly in handling the linguistic complexity of Arabic and the dialect variations present in tweets. Future work should expand the dataset to include other social media platforms and optimise the ML models, especially for Arabic text. Exploring advanced DL techniques such as transformer neural networks (TNNs) or RNNs could improve sentiment classification performance.

In conclusion, **MASHA** has demonstrated its potential as an effective solution for integrating ML models in SA for healthcare applications. Its modular design, scalability and ability to handle large-scale data make it a valuable framework for real-time SA in healthcare and beyond. Future enhancements, such as incorporating state-of-the-art ML models, including transformer-based architectures and ensemble DL approaches, can further enhance the framework’s predictive accuracy and robustness in public health monitoring. Additionally, expanding its application to other health-related domains, such as chronic disease management and mental health analysis, will demonstrate its versatility in addressing diverse healthcare challenges. Beyond healthcare, the framework’s modular and adaptable design makes it well-suited for adoption in other sectors, such as finance and cybersecurity, where real-time SA and data integration are pivotal for timely and data-driven decision-making.

## Data Availability

The dataset of Arabic tweets used in this study is not publicly available due to Twitter/X platform terms of service restrictions. The data may be made available upon reasonable request to the corresponding author, subject to applicable platform policies.

## Acknowledgments section

The author would like to acknowledge the annotators who contributed to the manual labelling of the Arabic tweet dataset used in this study. The author also extends gratitude to colleagues and reviewers whose constructive feedback helped improve this work. This research was conducted independently, without institutional or external financial support.

## Author contributions

Author contributions: S.Y.B.: Conceptualisation, Methodology, Software, Formal Analysis, Data Curation, Writing (Original Draft), Writing (Review & Editing).

## Declaration of generative AI and AI-assisted technologies in the writing process

During the preparation of this work, the author(s) used ***Grammarly*** in order to make grammar and spelling corrections and improve the sentence structure. After using this tool/service, the author(s) reviewed and edited the content as needed and take(s) full responsibility for the content of the publication.

